# Quantifying and characterizing hourly human exposure to malaria vectors bites in rural southwest Burkina Faso

**DOI:** 10.1101/2019.12.17.19014845

**Authors:** D.D Soma, B Zogo, P Taconet, A Somé, S Coulibaly, L Baba-Moussa, G.A Ouédraogo, A Koffi, C Pennetier, K.R Dabiré, N Moiroux

## Abstract

**Background:** To sustain the efficacy of malaria vector control, the World Health Organization (WHO) recommends the combination of effective tools. Before designing and implementing additional strategies in any setting, it is critical to monitor or predict when and where transmission occurs. However, to date, very few studies have quantified the behavioural interactions between humans and *Anopheles* vectors. Here, we characterized residual transmission in a rural area of Burkina Faso where long lasting insecticidal nets (LLIN) are widely used.

**Methods:** We analysed data on both human and malaria vectors behaviours from 27 villages to measure hourly human exposure to vector bites in dry and rainy seasons using mathematical models. We estimated the protective efficacy of LLINs and characterised where (indoors *vs*. outdoors) and when both LLIN users and non-users were exposed to vector bites.

**Results:** The percentage of the population who declared sleeping under a LLIN the previous night was very high regardless of the season, with an average LLIN use ranging from 92.43% to 99.89%. The use of LLIN provided > 80% protection against exposure to vector bites. The proportion of exposure for LLIN users was 29-57% after 05:00 and 0.05-12 % before 20:00. More than 80% of exposure occurred indoors for LLIN users and the estimate reached 90% for children under five years old in the dry cold season.

**Conclusions:** This study supports the current use of LLIN as a primary malaria vector control tool. It also emphasises the need to complement LLIN with indoor-implemented measures such as indoor residual spraying (IRS) and/or house improvement to effectively combat malaria in the rural area of Diébougou. Furthermore, malaria elimination programmes would also require strategies that target outdoor biting vectors to be successful in the area.

## Background

Massive distribution of long-lasting insecticidal nets (LLINs) is a core intervention for malaria control in Burkina Faso. Scaling-up of coverage with LLIN in sub-Saharan Africa has been very successful between 2000 and 2015 during which malaria morbidity and mortality have dropped considerably [1]. Unfortunately, this significant progress is stalling or even reversing in some countries. Burkina Faso is indeed one of the sixteen (16) in the world that documented an increase in malaria burden from 2016 to 2017 [2]. This trend might be attributed to the recent increases in prevalence and strength of pyrethroid resistance in malaria vectors [3–5]. Another possible cause is the development of behavioural resistance in vector populations [6–8]. In sub-Saharan Africa, there have been many reports of changes in vector species and/or vector biting behaviours to avoid contact with LLIN [6–8]. Such changes in vector populations are considered by many specialists as an important threat for indoor control strategies such as LLIN [9, 10].

To sustain the efficacy of vector control, the WHO recommends the combination of effective tools [11]. At present, there are a number of recommended tools available and many others under development that can potentially be combined with LLIN [12, 13]. However, national malaria control programs (NMCPs) are now facing challenges to design effective control strategies due to high variations in malaria epidemiology between and even within countries [14]. To do so, NMCP must be able to monitor or predict when and where transmission occurs. Entomological data alone are not sufficient to address this question, missing information about behaviours of local human populations in order to measure/predict human exposure to malaria vectors bite [15–17]. Unfortunately, only few studies to date have quantified and characterized human exposure to malaria vectors bites by analysing data on both human and vector behaviours [17].

The present work is a baseline study conducted in the Diébougou area, southwest Burkina Faso, to quantify the behavioural interactions between humans and *Anopheles* mosquitoes. Results of the entomological surveys previously reported [18] were used in combination with human behavioral data to study human exposure to *Anopheles* vector bites. The work is part of a large randomized control trial designed to investigate whether the combination of LLINs with other vector control tools can provide additional protection over malaria cases and transmission. The trial was carried out in Southwest Burkina Faso where malaria vectors shows high levels of pyrethroid resistance.

## Methods

This study was conducted in 27 villages located in the Diébougou health district, southwest Burkina Faso in order to collect baseline data for a randomized controlled trial (Fig. 1). These villages were selected based on geographical (distance between two villages higher than 2 km and accessibility during the rainy season) and demographic (a population size ranging from 200 to 500 inhabitants) criteria [18]. The climate in the study area is tropical with one dry season from October to April (including a cold period from December to February and a hot period from March to April) and one rainy season from May to September. Average daily temperature amplitudes are 18-36°C, 25-39°C and 23-33°C in dry cold, dry hot and rainy season, respectively. The mean annual rainfall is 1200 mm. The natural vegetation is dominated by wooded savannah dotted with clear forest gallery. The main economic activity is agriculture (cotton growing and cereals) followed by artisanal gold mining and production of coal and wood [19, 20].

**Fig 1.**
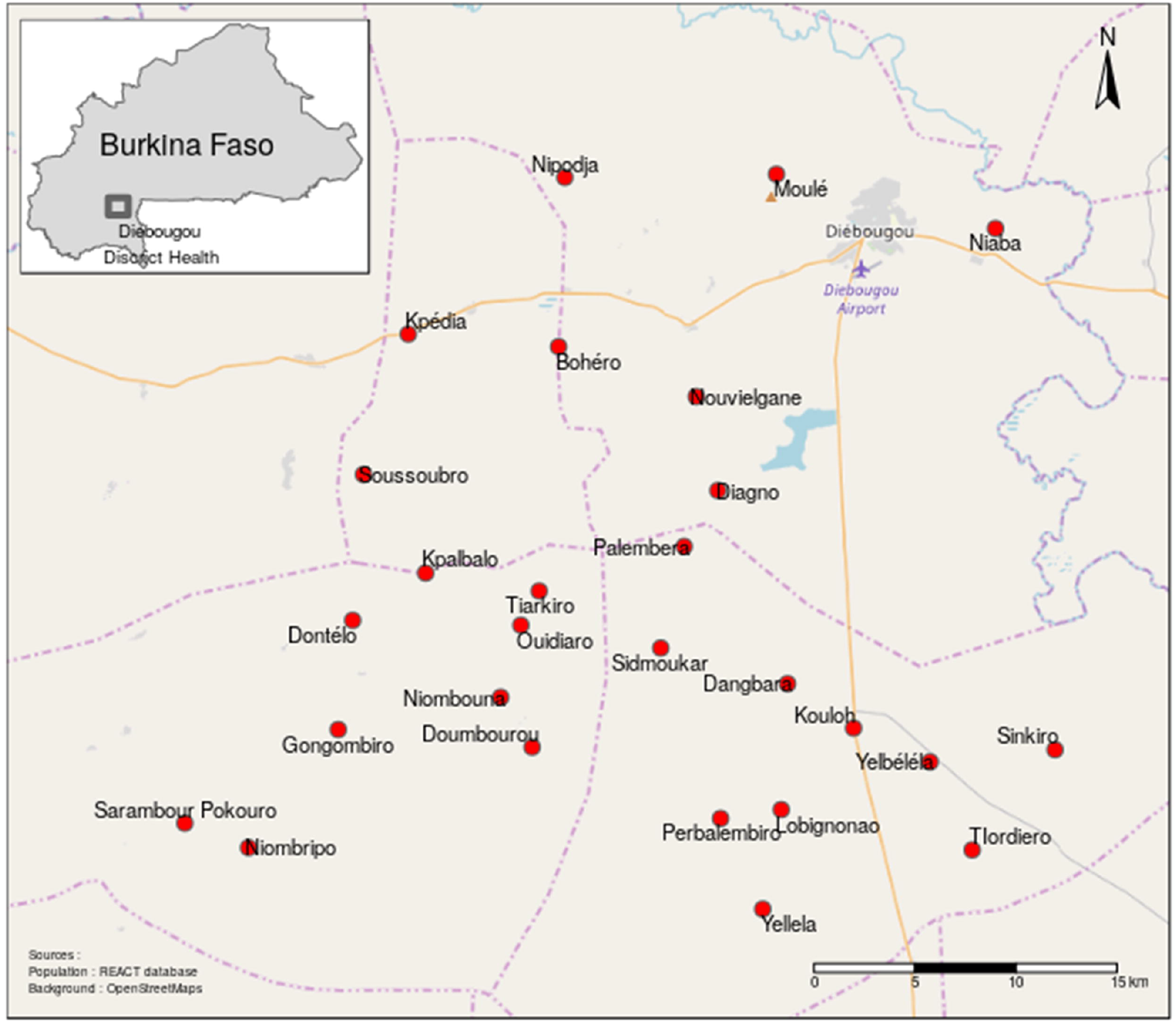
Map of the study area and villages surveyed.

We conducted three entomological surveys in the dry cold (January 2017), dry hot (March 2017) and rainy seasons (June 2017), respectively. During each survey, we collected human landing mosquitoes both indoors and outdoors from 17:00 h to 09:00 h in 4 houses per villages during one night [18]. All the mosquitoes collected were morphologically identified [21,22] and *Anopheles spp*. mosquitoes were subsequently identified to the species level by polymerase chain reaction [23–25]. Detailed descriptions of the methods used are provided in our previous publication [18]. Overall, *Anopheles funestus s*.*s* was the main malaria vector in the study area during the dry cold season [18]. During the dry hot and rainy seasons, *Anopheles coluzzii* and *Anopheles gambiae s*.*s* were the dominant species. The mean endophagy rate (ER) of malaria vectors was 63.23%, 50.18% and 57.18% during the dry cold, dry hot and rainy seasons, respectively [18].

In order to obtain appropriate data regarding relevant human behaviours, we surveyed 401 and 339 randomly selected households in dry (February to April 2017) and rainy (September 2017) seasons, respectively (corresponding to an average of 15 and 13 households per village). Among people usually leaving in each selected household, we randomly selected 3 persons (maximum) belonging to each of the 3 following age groups: 0-5 years old, 6-17 years old and ≥ 18 years old. We asked the head of the household the time at which each selected person (1) entered and exited his own house the night preceding the survey and (2) the time each LLIN user entered and exited his sleeping space the night preceding the survey [16]. In order to know the relative weight of each age group in the population, we recorded the number of individuals belonging to these groups in each households. A total of 3045 and 2880 individuals were surveyed representing 35.08% and 33.17 % of the 27 villages’ population according to a census carried out by our team in 2016 [18]. The human behavioural surveys were carried out using tablets running Open Data Kit (ODK) forms.

We used data from the human and *Anopheles* spp behavioural surveys to measure the human exposure to *Anopheles* spp. bites in dry season (cold and hot) and rainy season using mathematical models as previously described in Killeen et al. [15] and Moiroux et al. [16].

We estimated the average *true* personal protection of using a LLIN (i.e. the proportion of exposure to all bites occurring both indoors and outdoors that is prevented by using a LLIN) as well as the proportion of exposure which occurred indoors for LLIN users either accounting for the personal protection provided by net use or ignoring it to compare with available estimates for unprotected people. Exposure when sleeping under a LLIN was assumed to be reduced by 92% [16]. Moreover, to characterize residual transmission, we calculated the proportion of exposure occurring before 20:00 and after 5:00 (i.e. the times preceding and following the period when most (>50%) of LLIN users are protected).

All the exposure values were calculated at the village and study area levels, for each age group as well as for the total population. Because the number of individuals sampled per age group in each household is the same, the relative proportions of each age group in our sample are equal and do not reflect the relative proportions in the population. We therefore predicted the average number of people being indoors, outdoors and under nets at the population levels by summing weighted numbers of people of each age group. For these calculation and to produce figures, we used an R [26] package named “biteExp” developed by our team.

## Results

The average declared LLIN use rate was very high in the study population ranging from 95.49% in the dry season to 99.67% in the rainy season (Table 1). The declared LLIN use rate was higher in the 0-5 years old age group (97.87% in the dry season to 100% in the rainy season) compared to children aged 6-17 years old (95.36% in the dry season to 99.79% in the rainy season) and adults (92.45% in the dry season to 99.19% in the rainy season) (Table 1). However, we found that the LLIN use rate varied among villages (see Additional file 1) with the lowest rates observed in Kpédia (68.42%), Palembera (71.73%) and Diagnon (78.78%) in the adults group during the dry season. In the other villages, during dry and rainy seasons, LLIN use rates ranged from 80 to 100% whatever the season (see Additional file 1). Figure 2 shows humans and *Anopheles* behavior profiles as well as average hourly exposure and prevented exposure to bites for LLIN users in our study area.

**Table 1.** Average LLIN use rates, true average protection efficacy of LLINs against exposure to vector bites and proportions of indoors, “before bed” and “after bed” exposure to *Anopheles* bites for both LLIN users and non-users in 27 villages of the Diébougou area, Burkina Faso

| Season | Age<br>(years) | LLIN use rate<br>(%[min-max]) | *True average LLIN<br>personal protection<br>efficacy (% [min-max]) | Exposure indoors (%[min-max]) |  | Exposure before 20:00h<br>(%[min-max]) |  | Exposure after 05:00h<br>(%[min-max]) |  |
| --- | --- | --- | --- | --- | --- | --- | --- | --- | --- |
|  |  |  |  | LLIN users | Non-users | LLIN users | Non-users | LLIN users | Non-users |
| Dry cold<br>season | 18+ | 92.45 [68-100] | 83.44 [0-92] | 79.92 [0-100] | 96.67 [0-100] | 0.07 [0-0.13] | 0.04 [0-0.34] | 44.99 [0-100] | 8.16 [0-100] |
|  | 6 to 17 | 95.36 [71-100] | 83.79 [0-92] | 85.44 [0-100] | 97.64 [0-100] | 0.58 [0-1] | 0.12 [0-0.73] | 48.93 [0-100] | 9.01 [0-100] |
|  | 0 to 5 | 97.87 [81-100] | 86.73 [0-92] | 90.52 [0-100] | 98.74 [0-100] | 3.93 [0-100] | 0.62 [0-100] | 40.23 [0-100] | 12.20 [0-100] |
|  | population | 95.49 [77-100] | 84.93 [0-92] | 85.62 [0-100] | 97.83 [0-100] | 1.66 [0-100] | 0.31 [0-100] | 44.50 [0-100] | 10.11 [0-100] |
| Dry hot<br>season | 18+ | 92.45 [68-100] | 78.00 [0-92] | 69.57 [19-100] | 93.31 [75-100] | 3.38 [0-26] | 0.82 [0-1] | 57.20 [0-100] | 13.19 [0-100] |
|  | 6 to 17 | 95.36 [71-100] | 79.88 [2-92] | 82.70 [21-100] | 96.52 [72-100] | 4.57 [0-5] | 0.99 [0-2] | 56.20 [0-100] | 12.27 [0-100] |
|  | 0 to 5 | 97.87 [81-100] | 83.63 [13-92] | 88.73 [29-100] | 98.15 [82-100] | 11.30 [0-20] | 2.13 [0-3] | 43.95 [0-100] | 12.32 [0-100] |
|  | population | 95.49 [77-100] | 80.89 [5-92] | 80.54 [24-100] | 96.28 [78-100] | 6.56 [0-30] | 1.41 [0-2] | 52.19 [0-100] | 12.55 [0-100] |
| Rainy<br>season | 18+ | 99.19 [92-100] | 79.13 [53-92] | 75.61 [11-100] | 94.91 [62-100] | 10.08 [0-23] | 2.17 [0-5] | 42.90 [0-90] | 9.81 [0-44] |
|  | 6 to 17 | 99.79 [94-100] | 81.83 [51-92] | 83.28 [45-100] | 96.96 [91-100] | 10.24 [0-25] | 2.22 [0-8] | 48.59 [0-91] | 10.42 [0-50] |
|  | 0 to 5 | 100.00 | 87.00 [72-92] | 89.21 [69-100] | 98.60 [96-100] | 11.33 [0-19] | 2.31 [0-11] | 33.88 [0-85] | 10.55 [0-50] |
|  | population | 99.67 [97-100] | 82.82 [58-92] | 81.93 [27-100] | 96.90 [82-100] | 10.47 [0-23] | 2.23 [0-9] | 42.40 [0-89] | 10.27 [0-48] |

Min and max reported in brackets give the value recorded in the village with the lower and the higher average value, respectively.
\*True average LLIN personal protection efficacy: estimated proportion of *Anopheles* bites prevented by the use of a LLIN.

**Fig. 2.**
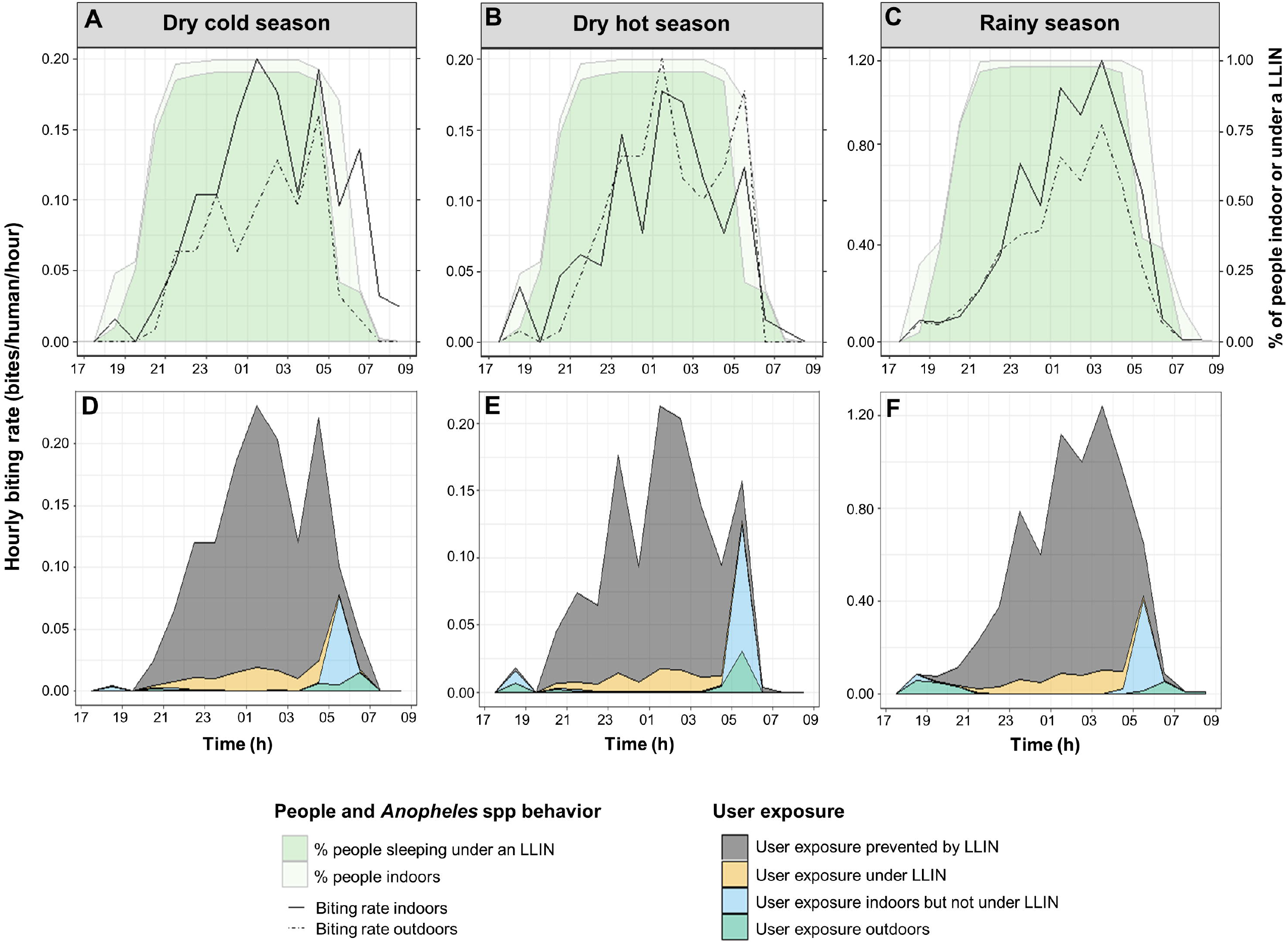
Hourly human and *Anopheles* spp behavior (A, B, C) and hourly exposure to bites of LLIN users (D, E, F) in the Diébougou health District, Burkina Faso. *Human behavioural data plotted in panel A and B are the same (only one dry season survey) but plotted with different entomological data*.

The majority of the population was indoors from 20:00 in both dry and rainy seasons (Figs. 2A, 2B and 2C). These populations woke up around 05:00 in the early morning in all seasons (Figs. 2A, 2B and 2C). Most of the total exposure to *Anopheles* bites occurred indoors (> 94%, Table 1) but was largely preventable by using of LLIN (Figs. 2D, 2E and 2F). Indeed, LLIN were estimated to provide average ‘true’ personal protection against 84.93%, 80.89% and 82.82% of exposure in dry cold season, dry hot season and rainy season, respectively (Table 1, Additional file 2). The peak of exposure for users occurred indoors between 05:00 and 06:00 just before sunrise whatever the season (Figs.2D, 2E and 2F). On average, between 33 and 57% of residual exposure of LLIN users occurred after wake up (after 5:00) depending on age groups. Early bites (before 20:00) represented less than 12 % of the residual exposure of LLIN users (Table 1).

## Discussion

The average declared LLIN use rate was very high (>95%) in all age groups of our study population. The LLIN use rate was slightly higher in children under five years of age than the rest of the population. This finding is consistent with results from a multi-country analysis that revealed that the most vulnerable groups are preferentially protected by LLIN in sub-Saharan Africa [27]. At the village level, the use rate rarely fall under 80%, being consistently higher than the nationwide LLIN use value of 67% published by WHO in 2017 [28]. This may be explained by the fact that the study was conducted approximately 6 months after a wide LLIN distribution. However, our reported LLIN use may be overestimated because it was based on self-reported survey questions, the most commonly used method to assess bednet use [29]. To more accurately estimate LLIN use, future studies quantifying human exposure to mosquito bites should consider using other measurement methods such as electronic monitoring devices [30, 31].

This study shows that the overall protective efficacy of LLINs against vector bites in the rural area of Diébougou was high (80-85 %) during the three seasons. Our estimates for LLIN personal efficacy were comparable with those found in Benin (80% and 87%) [16] but were higher than those reported elsewhere such as in Kenya (51 %) [32] and Tanzania (70%, 59% and 38%) [15, 33]. Our results support strongly the use of LLIN as a primary malaria vector control tool in the area. Nevertheless, such a protection level (85% in average) has to be put into perspective with the high malaria transmission and endemicity [18] in order to measure/realize the importance of malaria residual transmission in the area.

We estimated that 33-57% of residual exposure to *Anopheles* bites of LLIN users occurred after 5:00 and 0.07-12% occurred before 20:00 when most of users are awake. The proportion of exposure for LLIN users has been higher in the late part of the morning than in the early part of the evening in some settings while the opposite trend has been observed in other settings [15, 16, 34, 35]. In our study area, over 80% of human exposure to vector bites occurred indoors for LLIN users. For children under five years who use LLINs, the exposure rate occurring indoors reached 90%. Therefore, these results suggest that adding other indoor intervention such as indoor residual spraying (IRS) to LLINs would be relevant to reduce malaria transmission in the rural area of Diébougou. In 2017, 28 countries in the world have implemented IRS in combination with LLINs to combat malaria [2]. IRS contributed to an estimated 10 (5–14)% of the reduction in malaria burden achieved recently [36]. When used together, IRS and LLINs are expected to target vectors at different stages of their gonotrophic cycle using insecticides with different mode of action. However, trials assessing the impact of the combination IRS+LLIN over LLIN use alone have yielded conflicting results [37–42]. House improvement is another indoor measure which needs careful consideration and deep investigations. Indeed, house improvement has been strongly associated with reduced malaria transmission and disease in many studies [44–46]. The main house improvement interventions studied are closed eaves, closed ceilings, window screens and metal-roof houses as opposed to eaves, ceilings, windows openings and thatched-roof houses. Such improvements protect against malaria by providing physical barriers that prevent vectors from entering houses and can reduce vector survivorship [44, 47]. Nonetheless, there is compelling evidence that even a full coverage of effective measures within houses would not be sufficient to suppress transmission of malaria in Africa [43].

In this study, we evidenced that a significant proportion of LLIN users exposure to vector bites occurred outdoors (ranging from 9.48% to 30.43%), with the highest estimate recorded in adults (over the age of 18 years old) during the dry hot season. Many studies conducted in various areas of Africa reported similar or even higher estimates of exposure occurring outdoors [15, 33, 35, 48]. Recently, a systematic review categorized Burkina Faso along with Eritrea, Ethiopia, Gabon, and Tanzania as countries with high levels of outdoor vector biting [10]. However, our results do not fully support this categorization since we show that both LLIN users and LLIN non users are far more exposed to vector bites indoors than outdoors in the study area. Nevertheless, strategies targeting outdoor bites would probably be required to achieve malaria elimination in the area.

Almost all the existing indoor vector control strategies face two important evolutive challenges. First, they induce a strong selective pressure on physiological resistance in vector populations because they almost all rely on synthetic chemicals [49]. Second, they also induced selective pressure for behavioral changes in vector populations resulting in a reduced contact with interventions [49]. In this context, there is a crucial need to monitor these resistance mechanisms, as well as residual transmission, after the deployment of strategies to inform decision makers in order to allow them to adapt their strategic plans.

## Conclusions

This study showed that most of the population of the rural area of Diébougou reported using LLINs the previous night. The use of LLIN prevented more than 80% of *Anopheles* bite exposure. Nevertheless, LLIN users are still exposed to vector bites which occurred mostly indoors in late morning. Therefore, complementary strategies that target indoor biting vectors in combination with LLIN should be prioritized to control malaria in this area. However, as vectors are able to behaviourally evolve in response to indoor control tool implementation, successful malaria control programmes should also integrate monitoring of malaria vector behaviours. Moreover, as it is predictable that outdoor biting phenotypes will be selected, it urges to also evaluate and implement outdoor measures.

## Data Availability

The datasets used and/or analyzed during the current study are available from the corresponding author on reasonable request.

## List of abbreviations

(ER): Endophagy Rate
IRS: Indoor Residual Spraying
LLIN: Long-Lasting Insecticidal Nets
ODK: Open Data Kit
NMCP: National Malaria Control Programs
WHO: World Health Organization

## Declarations

### Ethics approval and consent to participate

The protocol of this study was reviewed and approved by the Institutional Ethics Committee of the Institut de Recherche en Sciences de la Santé (IEC-IRSS) and registered as N°A06/2016/CEIRES. We received community agreement before the beginning of human and *Anopheles* spp behavioral surveys. Behavioral surveys did not involved participants under 16 years old. Indeed, questionnaires were administrated only to the heads of households and informations relative to children under 16 years old were therefore directly collected from either a parent or guardian. All mosquito collectors involved in the entomological surveys gave their informed consent but none of them were under 16 years old. Mosquito collectors and supervisors received a vaccine against yellow fever as a prophylactic measure. Collectors were treated free of charge for malaria according to WHO recommendations.

### Consent for publication

Not applicable.

### Authors’ contributions

NM, RKD and DDS conceived and designed the study. DDS and SC collected the data. DDS and NM analyzed the data. DDS and BZ drafted the manuscript. NM, CP, PT, AS, LMB, GAO, AK and RKD reviewed the manuscript; all authors read and approved the final manuscript.

## Acknowledgements

We acknowledge the Burkina Faso Ministry of Health, particularly Dr. Dembélé Henri and local medical team who facilitated the data collection. We thank all the villagers and local authorities for their kind collaboration throughout the study. Special thanks are due to Mr. Maiga Issouf for his strong commitment during human behavioural surveys. We are very grateful to Mr. Dahounto Amal for their substantial contributions and collaboration. We thank all the IRSS field staff for their assistance; the “Laboratoire Mixte International sur les Maladies à Vecteurs” (LAMIVECT) for their technical support. We also thank to Mr. Ouattara Adama for their support.

## Figures, tables and additional files

**Additional file 1**. LLIN Use rate per village

**Additional file 2**. True average protection efficacy of LLINs against transmission and Proportions of indoors, early evening and late morning exposure to *Anopheles* bites per village

## References

1. Bhatt S, Weiss DJ, Cameron E, Bisanzio D, Mappin B, Dalrymple U, et al. The effect of malaria control on Plasmodium falciparum in Africa between 2000 and 2015. Nature. 2015;526:207–11.

2. WHO. World malaria report 2018. 2018;238. http://www.who.int/malaria/publications/world-malaria-report-2018.

3. Dabiré KR, Diabaté A, Djogbenou L, Ouari A, N’Guessan R, Ouédraogo J-B, et al. Dynamics of multiple insecticide resistance in the malaria vector Anopheles gambiae in a rice growing area in South-Western Burkina Faso. Malar J. 2008;7:188.

4. Toé KH, Jones CM, N’fale S, Ismai HM, Dabiré RK, Ranson H. Increased pyrethroid resistance in malaria vectors and decreased bed net effectiveness Burkina Faso. Emerg Infect Dis. 2014;20:1691–6.

5. Toé KH, N’Falé S, Dabiré RK, Ranson H, Jones CM. The recent escalation in strength of pyrethroid resistance in Anopheles coluzzi in West Africa is linked to increased expression of multiple gene families. BMC Genomics. 2015;16:1–11.

6. Ojuka P, Boum Y, Denoeud-Ndam L, Nabasumba C, Muller Y, Okia M, et al. Early biting and insecticide resistance in the malaria vector Anopheles might compromise the effectiveness of vector control intervention in Southwestern Uganda. Malar J. 2015;14:1–8.

7. Moiroux N, Gomez MB, Pennetier C, Elanga E, Djènontin A, Chandre F, et al. Changes in anopheles funestus biting behavior following universal coverage of long-lasting insecticidal nets in benin. J Infect Dis. 2012;206:1622–9.

8. Fornadel CM, Norris LC, Glass GE, Norris DE. Analysis of Anopheles arabiensis blood feeding behavior in southern zambia during the two years after introduction of insecticide-treated bed nets. Am J Trop Med Hyg. 2010;83:848–53.

9. Ranson H, Lissenden N. Insecticide Resistance in African Anopheles Mosquitoes: A Worsening Situation that Needs Urgent Action to Maintain Malaria Control. Trends Parasitol. 2016;32:187–96. doi:10.1016/j.pt.2015.11.010.

10. Sherrard-Smith E, Skarp JE, Beale AD, Fornadel C, Norris LC, Moore SJ, et al. Mosquito feeding behavior and how it influences residual malaria transmission across Africa. Proc Natl Acad Sci U S A. 2019;:201820646.

11. WHO. Global plan for insecticide resistance management in malaria vectors. World Health Organization. 2012;:13. https://apps.who.int/iris/bitstream/handle/10665/44846/9789241564472_eng.pdf.

12. Killeen GF, Tatarsky A, Diabate A, Chaccour CJ, Marshall JM, Okumu FO, et al. Developing an expanded vector control toolbox for malaria elimination. BMJ Glob Heal. 2017;2:e000211.

13. Barreaux P, Barreaux AMG, Sternberg ED, Suh E, Waite JL, Whitehead SA, et al. Priorities for Broadening the Malaria Vector Control Tool Kit. Trends Parasitol. 2017;33:763–74.

14. Kelly-Hope LA, McKenzie FE. The multiplicity of malaria transmission: A review of entomological inoculation rate measurements and methods across sub-Saharan Africa. Malaria Journal. 2009;8:19.

15. Killeen GF, Kihonda J, Lyimo E, Oketch FR, Kotas ME, Mathenge E, et al. Quantifying behavioural interactions between humans and mosquitoeslJ: Evaluating the protective efficacy of insecticidal nets against malaria transmission in rural Tanzania. BMC Infect Dis. 2006;6:1–10.

16. Moiroux N, Damien GB, Egrot M, Djenontin A, Chandre F, Corbel V, et al. Human exposure to early morning Anopheles funestus biting behavior and personal protection provided by long-lasting insecticidal nets. PLoS One. 2014;9:8–11.

17. Monroe A, Moore S, Koenker H, Lynch M, Ricotta E. Measuring and characterizing night time human behaviour as it relates to residual malaria transmission in sub - Saharan AfricalJ: a review of the published literature. Malar J. 2019;:1–12. doi:10.1186/s12936-019-2638-9.

18. Soma DD, Zogo BM, Somé A, Tchiekoi BN, Hien DF de S, Pooda HS, et al. Anopheles bionomic, insecticide resistance and malaria transmission in southwest Burkina FasolJ: a pre-intervention study. BioRxiv Prepr. 2019.

19. INSD. Tableau de bord économique et social 2014 de la région du Sud Ouest. 2015.

20. INSD. Enquête nationale sur le secteur de l’orpaillage (ENSO). 2017.

21. Mattingly PF, Rageau J, La CA, Des F, Du M, Xii SA. Contributions de la faune des moustiques du Sud-Est Asiatique. XII. Contrib Am Entomol Inst. 1973;7.

22. Gillies MT and Coetzee M. A supplement to the Anophelinae of Africa South of the Sahara (Afrotropical Region). South African Inst Med Res. 1987;:143.

23. Koekemoer LL, Kamau L, Hunt RH, Coetzee M. A cocktail polymerase chain reaction assay to identify members of the Anopheles funestus (Diptera: Culicidae) group. Am J Trop Med Hyg. 2002;66:804–11.

24. Cohuet A, Simard F, Berthomieu A, Raymond M, Fontenille D WM. Isolation and characterization of microsatellite DNA markers in the malaria vector Anopheles funestus. Mol Ecol Notes. 2002;2:498–500.

25. Santolamazza F, Mancini E, Simard F, Qi Y, Tu Z, della Torre A. Insertion polymorphisms of SINE200 retrotransposons within speciation islands of Anopheles gambiae molecular forms. Malar J. 2008;7:163. doi:10.1186/1475-2875-7-163.

26. The R Development Core Team. R□: A Language and Environment for Statistical Computing. 2008;2:1–2547. http://www.gnu.org/copyleft/gpl.html.

27. Olapeju B, Choiriyyah I, Lynch M, Acosta A, Blaufuss S, Filemyr E, et al. Age and gender trends in insecticide-treated net use in sub-Saharan Africa: a multi-country analysis. Malar J. 2018;17:423.

28. WHO. World Malaria Report 2017. 2017. doi:10.1071/EC12504.

29. Krezanoski PJ, Bangsberg DR, Tsai AC. Quantifying bias in measuring insecticide-treated bednet use: meta-analysis of self-reported vs objectively measured adherence. J Glob Health. 2018;8:010411.

30. Koudou BG, Malone D, Hemingway J. The use of motion detectors to estimate net usage by householders, in relation to mosquito density in central Cote d’Ivoire: Preliminary results. Parasites and Vectors. 2014.

31. Krezanoski PJ, Santorino D, Agaba A, Dorsey G, Bangsberg DR, Carroll RW. How Are Insecticide-Treated Bednets Used in Ugandan Households? A Comprehensive Characterization of Bednet Adherence Using a Remote Monitor. Am J Trop Med Hyg. 2019;00:1–8.

32. Cooke MK, Kahindi SC, Oriango RM, Owaga C, Ayoma E, Mabuka D, et al. ‘A bite before bed’: exposure to malaria vectors outside the times of net use in the highlands of western Kenya. Malar J. 2015;14:259.

33. Geissbühler Y, Chaki P, Emidi B, Govella NJ, Shirima R, Mayagaya V, et al. Interdependence of domestic malaria prevention measures and mosquito-human interactions in urban Dar es Salaam, Tanzania. Malar J. 2007;6:126.

34. Kamau A, Mwangangi JM, Rono MK, Mogeni P, Omedo I, Midega J, et al. Variation in the effectiveness of insecticide treated nets against malaria and outdoor biting by vectors in Kilifi, Kenya. Wellcome open Res. 2017;2:22.

35. Russell TL, Govella NJ, Azizi S, Drakeley CJ, Kachur SP, Killeen GF. Increased proportions of outdoor feeding among residual malaria vector populations following increased use of insecticide-treated nets in rural Tanzania. Malar J. 2011;10:80. doi:10.1186/1475-2875-10-80.

36. Bhatt S, Weiss DJ, Cameron E, Bisanzio D, Mappin B, Dalrymple U, et al. The effect of malaria control on Plasmodium falciparum in Africa between 2000 and 2015. Nature. 2015;526:207–11.

37. West PA, Protopopoff N, Wright A, Kivaju Z, Tigererwa R, Mosha FW, et al. Indoor Residual Spraying in Combination with Insecticide-Treated Nets Compared to Insecticide-Treated Nets Alone for Protection against MalarialJ: A Cluster Randomised Trial in Tanzania. PLOS Med. 2014;11:1–12.

38. Protopopoff N, Mosha JF, Lukole E, Charlwood JD, Wright A, Mwalimu CD, et al. Effectiveness of a long-lasting piperonyl butoxide-treated insecticidal net and indoor residual spray interventions, separately and together, against malaria transmitted by pyrethroid-resistant mosquitoes: a cluster, randomised controlled, two-by-two fact. Lancet. 2018;:1–12. doi:10.1016/S0140-6736(18)30427-6.

39. Kafy HT, Ismail BA, Mnzava AP, Lines J, Abdin MSE, Eltaher JS, et al. Impact of insecticide resistance in Anopheles arabiensis on malaria incidence and prevalence in Sudan and the costs of mitigation. Proc Natl Acad Sci. 2017;114:E11267–75.

40. Corbel V, Akogbeto M, Damien GB, Djenontin A, Chandre F, Rogier C, et al. Combination of malaria vector control interventions in pyrethroid resistance area in Benin: acluster randomised controlled trial. Lancet Infect Dis. 2012;12:617–26.

41. Pinder M, Jawara M, Jarju LBS, Salami K, Jeffries D, Adiamoh M, et al. Efficacy of indoor residual spraying with dichlorodiphenyltrichloroethane against malaria in Gambian communities with high usage of long-lasting insecticidal mosquito nets: A cluster-randomised controlled trial. Lancet. 2015;385:1436–46.

42. Loha E, Deressa W, Gari T, Balkew M, Kenea O, Solomon T, et al. Long-lasting insecticidal nets and indoor residual spraying may not be sufficient to eliminate malaria in a low malaria incidence area: results from a cluster randomized controlled trial in Ethiopia. Malar J. 2019;18:141.

43. World Health Organization. Control of residual malaria parasite transmission. 2014.

44. Tusting LS, Ippolito MM, Willey BA, Kleinschmidt I, Dorsey G, Gosling RD, et al. The evidence for improving housing to reduce malaria: A systematic review and meta-analysis. Malaria Journal. 2015.

45. Rek JC, Alegana V, Arinaitwe E, Cameron E, Kamya MR, Katureebe A, et al. Rapid improvements to rural Ugandan housing and their association with malaria from intense to reduced transmission: a cohort study. Lancet Planet Heal. 2018.

46. Killeen GF, Govella NJ, Mlacha YP, Chaki PP. Suppression of malaria vector densities and human infection prevalence associated with scale-up of mosquito-proofed housing in Dar es Salaam, Tanzania: re-analysis of an observational series of parasitological and entomological surveys. Lancet Planet Heal. 2019;3:e132–43.

47. Lindsay SW, Jawara M, Mwesigwa J, Achan J, Bayoh N, Bradley J, et al. Reduced mosquito survival in metal-roof houses may contribute to a decline in malaria transmission in sub-Saharan Africa. Sci Rep. 2019;9:7770.

48. Huho B, Briët O, Seyoum A, Sikaala C, Bayoh N, Gimnig J, et al. Consistently high estimates for the proportion of human exposure to malaria vector populations occurring indoors in rural Africa. Int J Epidemiol. 2013;42:235–47.

49. Lies Durnez and Marc Coosemans. Residual Transmission of Malaria: An Old Issue for New Approaches. In: Anopheles mosquitoes - New insights into malaria vectors. Intech Open Science; 2013. p. 671–704. doi:http://dx.doi.org/10.5772/55925.

